# Strengthening Rabies Surveillance Capacity Using Existing Resources through an Integrated Bite Case Management Model: A Practical One Health Action

**DOI:** 10.64898/2026.02.05.26345692

**Authors:** Do Muoi Le Thuong, Linh Thi My Phan, Anh The Dao, Thieu Hoang Le, Anh The Le, Trang Thi Thuy Vo, Anh Quang Dang, Linh Thi Thuy Do, Nghi Vinh Pham, Ha Thi Cam Pham, Huong Thi Thanh Nguyen, Hung Thai Do

**Author notes:** Corresponding author **Email:** **(TDML)**.

## Abstract

**Background:** Rabies remains a cause of mortality in many low- and middle-income countries, with the majority of human infections resulting from dog-to-human transmission. The Integrated Bite Case Management (IBCM) model is a One Health approach that aims to strengthen rabies surveillance and response by linking the management of human bite cases with investigation of the implicated animals. This study aimed to evaluate the effectiveness of implementing IBCM in Quang Nam Province under existing resource conditions.

**Methodology/Principal Findings:** A pre–post intervention study without a control group was conducted across the entire province. During the intervention period, 11,673 animal-bite cases were recorded; IBCM identified 75 animals suspected of having rabies, of which 40 tested positive for rabies virus by RT-PCR. Most of these animals were unvaccinated, free-roaming dogs. In communes where outbreaks were detected, the average number of registered dogs increased from 507 to 543 per commune, and vaccination coverage increased from 44.1% to 72.6% within 21 days. The average number of Post-exposure prophylaxis (PEP) courses administered per month increased from 349 to 971, the proportion of high-risk exposures increased from 9.3% to 11.9%, and the proportion of delayed PEP (≥10 days after exposure) rose slightly from 5.9% to 6.6%. At the same time, the proportion of staff with good knowledge of rabies diagnosis in animals increased substantially, from 9.1% to 55.6%. The main limitations included the pre–post design and loss to follow-up of some animals, which prevented laboratory testing.

**Conclusion:** The implementation of IBCM within the existing health and veterinary systems substantially strengthened rabies surveillance and response in accordance with the One Health approach. IBCM was demonstrated to be feasible, resource-appropriate, and scalable, thereby contributing to progress toward the global goal of eliminating human deaths from dog-mediated rabies by 2030.

**Author summary:** Rabies is a preventable disease, yet it continues to cause deaths in many countries where dogs remain the primary reservoir and source of infection. In Vietnam, rabies surveillance remains largely separated between the human and animal health sectors. The IBCM model uses human bite cases as the “trigger point” for coordinated investigation of the implicated animals, risk assessment, and information sharing between the two sectors, thereby supporting both clinical decision-making and outbreak response. We implemented the IBCM model in Quang Nam Province and observed an increase in the number of rabid animals detected, a marked rise in dog vaccination coverage in outbreak-affected areas, and substantial improvement in the knowledge and capacity of both health and veterinary staff. Simply by strengthening collaboration and information sharing between the two sectors, the rabies surveillance system became more sensitive and effective. This represents a practical example of One Health approach in action.

## Introduction

Rabies is a zoonotic infectious disease characterized by acute neurological progression and an almost invariably fatal outcome once clinical symptoms appear. Although effective preventive measures for rabies have been developed and standardized for decades, the disease still causes approximately 59,000 deaths globally each year [1]. With the exception of Antarctica, rabies remains endemic across almost all continents. Around 95% of rabies-related deaths occur in Asia and Africa, imposing a substantial health and economic burden on these regions [1, 2]. In Southeast Asia, rabies continues to show complex and concerning epidemiological trends and has one of the regions with the highest incidence compared with other parts of Asia [3]. Throughout the evolution of human understanding of rabies, as well as evidence from epidemiological studies, domestic dogs have consistently been identified as the primary source of human infection, accounting for approximately 99% of human rabies deaths [1, 4].

In this context, the World Health Organization (WHO), the World Organisation for Animal Health (WOAH, formerly OIE), and the Food and Agriculture Organization of the United Nations (FAO) jointly adopted the global strategy “Zero human deaths from dog-mediated rabies by 2030” (Zero by 30), which emphasizes the central role of mass dog vaccination, timely access to PEP, and strengthened rabies surveillance under a One Health approach [1]. Among these pillars, surveillance is considered a foundational component for early risk detection, guiding interventions, and optimizing resource usefor rabies prevention and control. However, in many endemic countries, rabies surveillance is poorly coordinated between the human and animal health sectors, resulting in incomplete reporting, delayed risk assessment, and challenges in linking information on human exposures with the rabies status of animals [5]. Consequently, a considerable proportion of PEP decisions are made in the absence of adequate information regarding the source of exposure, while many rabies cases in animals remain undetected or insufficiently investigated, thereby undermining the effectiveness of dog-focused rabies control strategies.

The IBCM model was developed to address these gaps by closely integrating human and animal health surveillance activities around human exposure events. At its core, IBCM uses information from individuals bitten by animals as the trigger for investigating suspected rabid animals, combining clinical and epidemiological risk assessment to support appropriate PEP decision-making, while simultaneously enhancing the detection of rabies cases in animals within the community [6]. Although promising, empirical evidence on the effectiveness of IBCM remains unevenly distributed, particularly with respect to studies evaluating its application under routine, resource-constrained local health system conditions.

Vietnam is among the countries with the highest reported mortality from rabies, with approximately 70–100 human deaths recorded annually, and rabies has remained one of the infectious diseases with the highest case fatality in the country for many years [7]. Rabies surveillance programs in Vietnam were established relatively early and have achieved substantial progress in reducing the burden of the disease; however, the system still faces several limitations, including shortages of human resources, uneven staff capacity, and low data quality [8, 9]. IBCM was first piloted in Vietnam in 2016 in Phu Tho Province with support from the United States Centers for Disease Control and Prevention (US CDC), and the model demonstrated effectiveness in detecting rabies cases in dogs [10]. In 2024, IBCM was implemented in Quang Nam Province, now under the administrative management of Da Nang City, where consistently low dog vaccination coverage has placed the population at high risk of rabies transmission. This report describes in detail the activities and outcomes of implementing the IBCM model using existing local resources, and compares the effectiveness of the model in improving rabies surveillance capacity with the period prior to IBCM implementation.

## Methods

The IBCM model was first introduced in Quang Nam Province at the end of August 2024 during a regional intersectoral training workshop on human–animal collaboration in rabies prevention and control in Central Vietnam, organized by the National Institute of Hygiene and Epidemiology. The training programme included: an introduction to IBCM and its operational framework; training in rabies diagnosis in animals for both health and veterinary personnel; establishment of regional IBCM groups comprising human and animal health staff via Zalo (a widely used group-messaging application in Vietnam); and development of standardized reporting procedures together with guidance on how animal investigation findings should be used to support outbreak response and PEP decision-making. Operational data for the IBCM model were recorded from September 2024 through August 2025 (12 months), after which the results were compared with the pre-intervention period. A final review workshop was held in October 2025 to evaluate IBCM performance and to provide direction for continued implementation.

### Study setting

The study was conducted in Quang Nam Province (Fig 1). Prior to 1 July 2025, Quang Nam consisted of 18 districts; from 1 July 2025, Vietnam implemented a two-tier local government model, under which Quang Nam Province was administratively merged into Da Nang City, and the Quang Nam area was reorganized into 78 communes/wards. The total population of this area was estimated at 1,539,468 according to the 2024 mid-term population census. As of the end of August 2024, the total dog population in the area was estimated at 110,332, although distribution was uneven, with higher concentrations in densely populated areas. Dog vaccination coverage during 2021–2023 remained below 20%.

**Fig 1.**
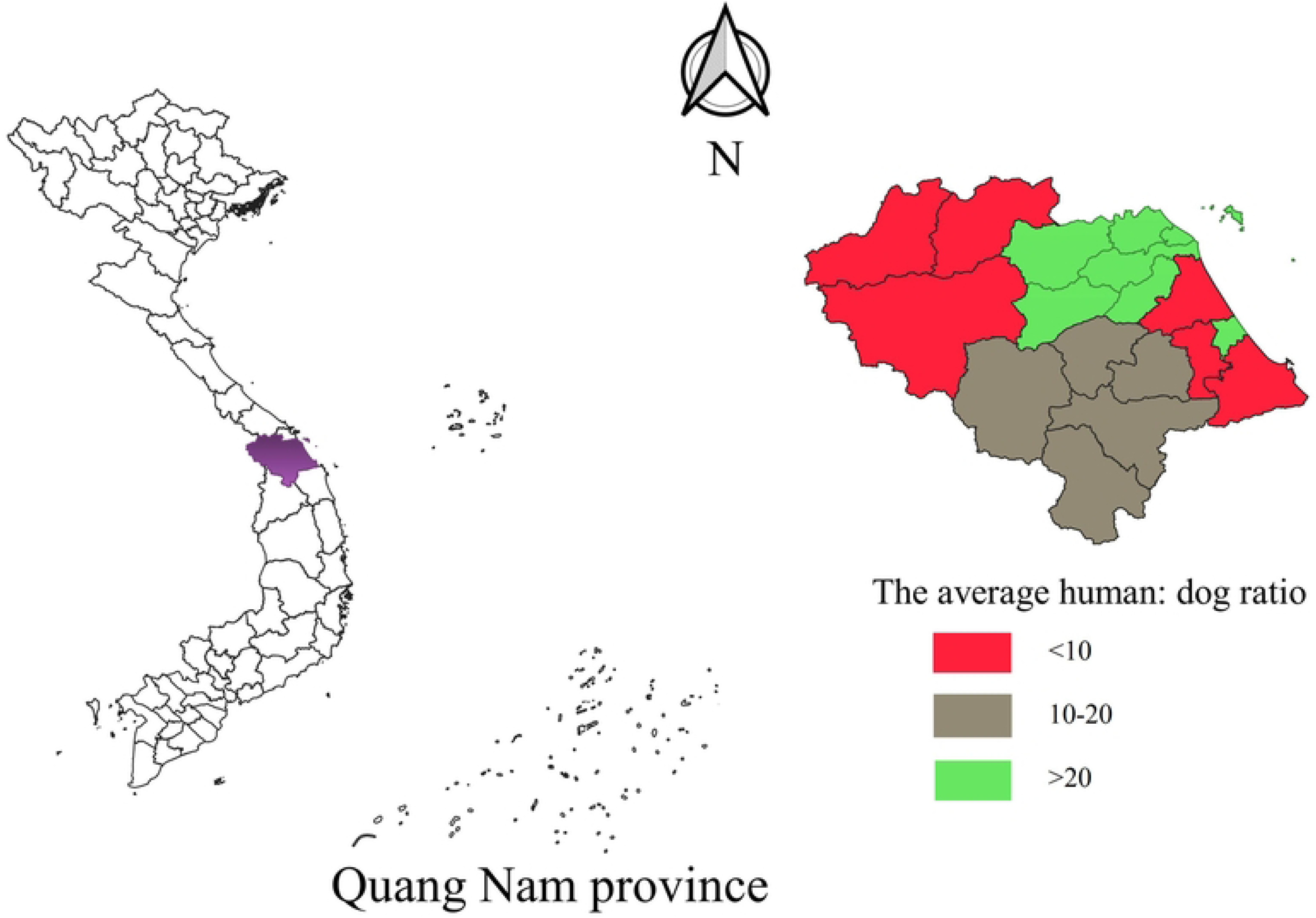
Study area and estimated human-to-dog ratios in Quang Nam Province Map showing the location of Quang Nam Province (purple), and estimated human-to-dog ratios across the study area. Areas shown in green indicate low dog density with a human-to-dog ratio greater than 20:1; areas in grey indicate moderate dog density with a ratio of 10–20:1; and areas in red indicate high dog density with a human-to-dog ratio of less than 10:1.

### Study population

All bite incidents occurring within the 12-month period following the introduction of the IBCM model, from 1 September 2024 to 30 August 2025, were recorded and classified. Animals suspected of rabies were investigated and, where feasible, sampled for laboratory testing based on the standardized case definition for rabies in animals. Information on suspected rabid animals was collected using a pre-designed IBCM investigation form. All health and veterinary personnel involved in rabies surveillance under the IBCM model were surveyed to assess their knowledge of rabies diagnosis in animals at two time points: prior to the intervention and 12 months after its completion.

### IBCM framewwork

IBCM was implemented through five key steps: (1) reporting high-risk bite incidents; (2) investigating information related to suspected rabid animals; (3) sharing information on suspected rabid animals between the human and animal health sectors; (4) conducting secondary investigations when required; and (5) using the collected information to guide appropriate public health and veterinary responses (Fig 2).

**Fig 2.**
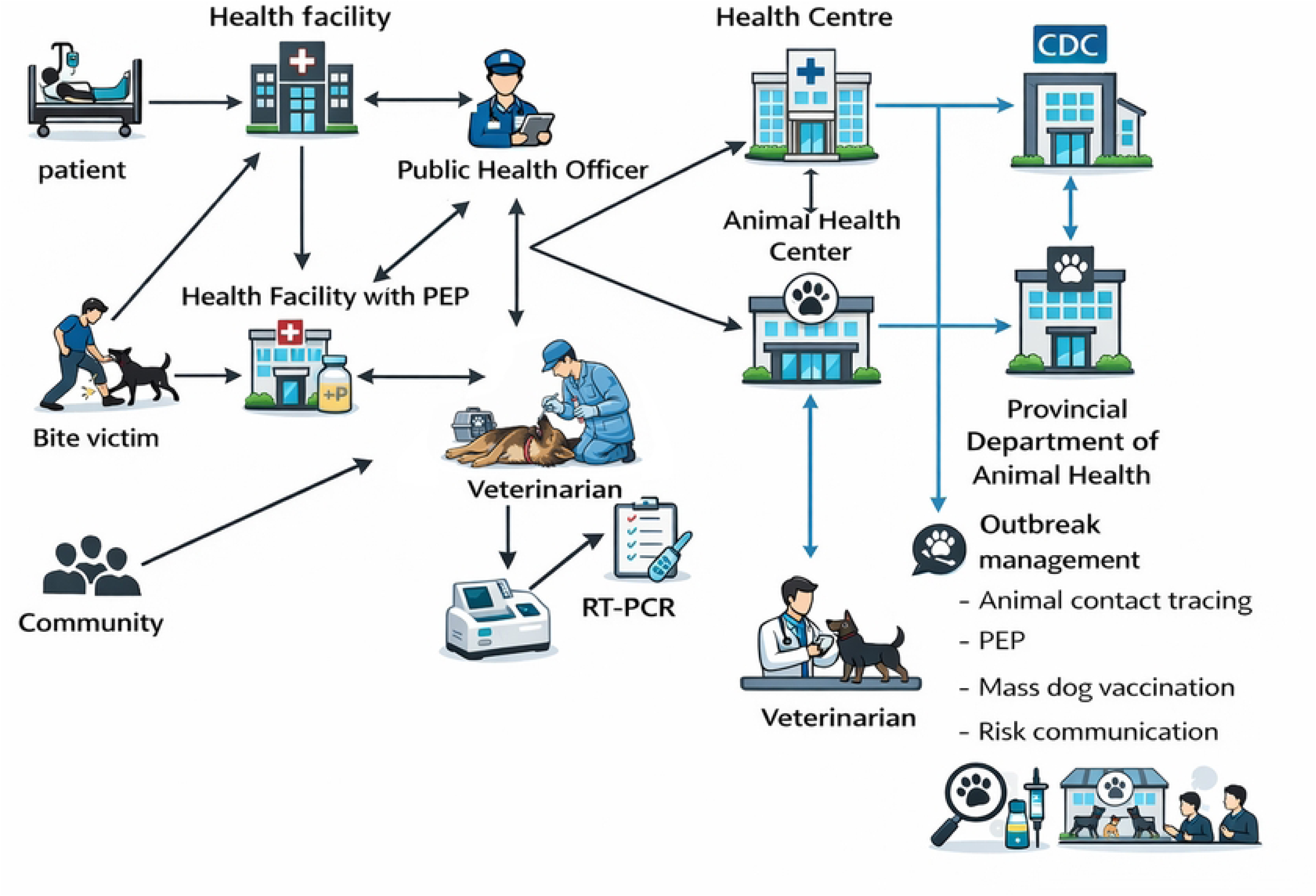
IBCM implementation framework in Quang Nam. Sources of reported bite incidents included PEP administration units, hospitals, and community reports. Suspected animals were screened and sampled by veterinary staff. A community health worker coordinated the programme and notified both the human and animal health sectors of detected cases to support joint decision-making on appropriate response measures.

### Criteria for triggering animal investigation and case definitions in animals

Not every animal bite automatically triggers the IBCM investigation process. To minimise the surveillance workload, risk classification of the biting animal was conducted promptly upon notification of a bite incident, based on predefined exclusion criteria and recognised clinical signs suggestive of rabies in animals [11]. If the animal was classified as being at risk of rabies based on its clinical presentation, veterinary staff collected specimens for laboratory testing and submitted them for RT-PCR analysis at the designated veterinary laboratory. The time interval from sample collection to the availability of test results typically ranged from two to three days. Laboratory confirmation was subsequently used to classify the level of rabies risk in the animal in accordance with WHO guidance.

### Animal rabies risk classification

− Low-risk animal: The biting animal shows no clinical signs consistent with rabies, remains healthy and alive for 10 days following the bite/exposure event, or tests negative for rabies.
− Undetermined risk: The biting animal cannot be identified or located; therefore, its history is unknown (e.g., vaccination or health status, exposure to suspected, probable, or confirmed rabid animals, or current clinical condition).
− High-risk animal: The biting animal presents with clinical signs suggestive of rabies (e.g., aggressive or abnormal behaviour, hypersalivation, paralysis, tremors, abnormal vocalisation, anorexia); and/or has a known history of exposure to a suspected or confirmed rabid animal; and dies within 10 days following the exposure event; or tests positive for rabies.

### Establishing IBCM using existing resources

From 2022 to August 2024, a rabies viral circulation surveillance programme funded by the U.S. Centers for Disease Control and Prevention (CDC) was implemented in Quang Nam Province. A total of 108 animal specimens were collected and tested using RT-PCR; however, all samples were negative for rabies virus. Although this circulation surveillance programme helped strengthen specimen collection and laboratory capacity among provincial veterinary staff, it did not demonstrate effectiveness in detecting rabid animals in the community.

Recognising the limitations of circulation surveillance, as well as the favourable conditions already in place, the IBCM model was introduced through an intersectoral training workshop for human and animal health staff at the end of August 2024. Health-care and veterinary workers were comprehensively trained in bite-risk classification and clinical diagnosis of rabies in animals based on WHO recommendations [11], enabling the early exclusion of animals with no rabies risk and thereby reducing the burden of field investigations.

Laboratory testing costs for animal samples were fully covered by the local government under pre-existing animal disease control policies. Human resources participating in IBCM activities were entirely drawn from the existing public health and veterinary systems, without the need to recruit additional staff or provide programme-specific financial incentives. Information exchange and rapid case notification were conducted via Zalo group messaging (a free and widely-used communication platform in Viet Nam), rather than adopting dedicated e-IBCM software, due to concerns regarding language barriers, licensing costs, and uncertainty about user acceptability among health-care and veterinary personnel.

### Data management and analysis

All IBCM forms were collected and cross-checked against official documents (including laboratory test result reports and administrative notices issued by local authorities). The validated data were then entered using Epidata software. Spatial data were analysed using QGIS version 3.22.0 to describe the geographic distribution of rabies-related risk and potential transmission chains. Descriptive statistical analyses, including frequencies, proportions, and rates, were performed using SPSS version 20.0. Intervention-period data were compared with pre-intervention data to assess changes in rabies surveillance performance following the implementation of IBCM.

### Ethics statement

This study was conducted to strengthen rabies surveillance capacity within the human and animal health sectors. Ethical approval was obtained from the Institutional Review Board of the National Institute of Hygiene and Epidemiology (Certificate No:NIHE IRB-19/2024; S1 Appendix). The study was implemented with the approval of local authorities and in close collaboration with the provincial public health and veterinary services in Quang Nam.

## Results

From 1 September 2024 to 30 August 2025, a total of 11,673 bite incidents were recorded (Fig 3). Fifty-one intersectoral investigations were triggered based on reports from PEP-providing vaccination units (n = 49) and hospitals (n = 2). From these health-sector reports, 35 animal specimens were collected for laboratory testing, of which 25 tested positive for rabies virus by RT-PCR and 10 tested negative. In addition, 24 investigations were initiated following bite incidents reported by the community. Among these, 21 animal samples met the IBCM sampling criteria, with 15 samples testing positive and 6 testing negative for rabies.

**Fig 3.**
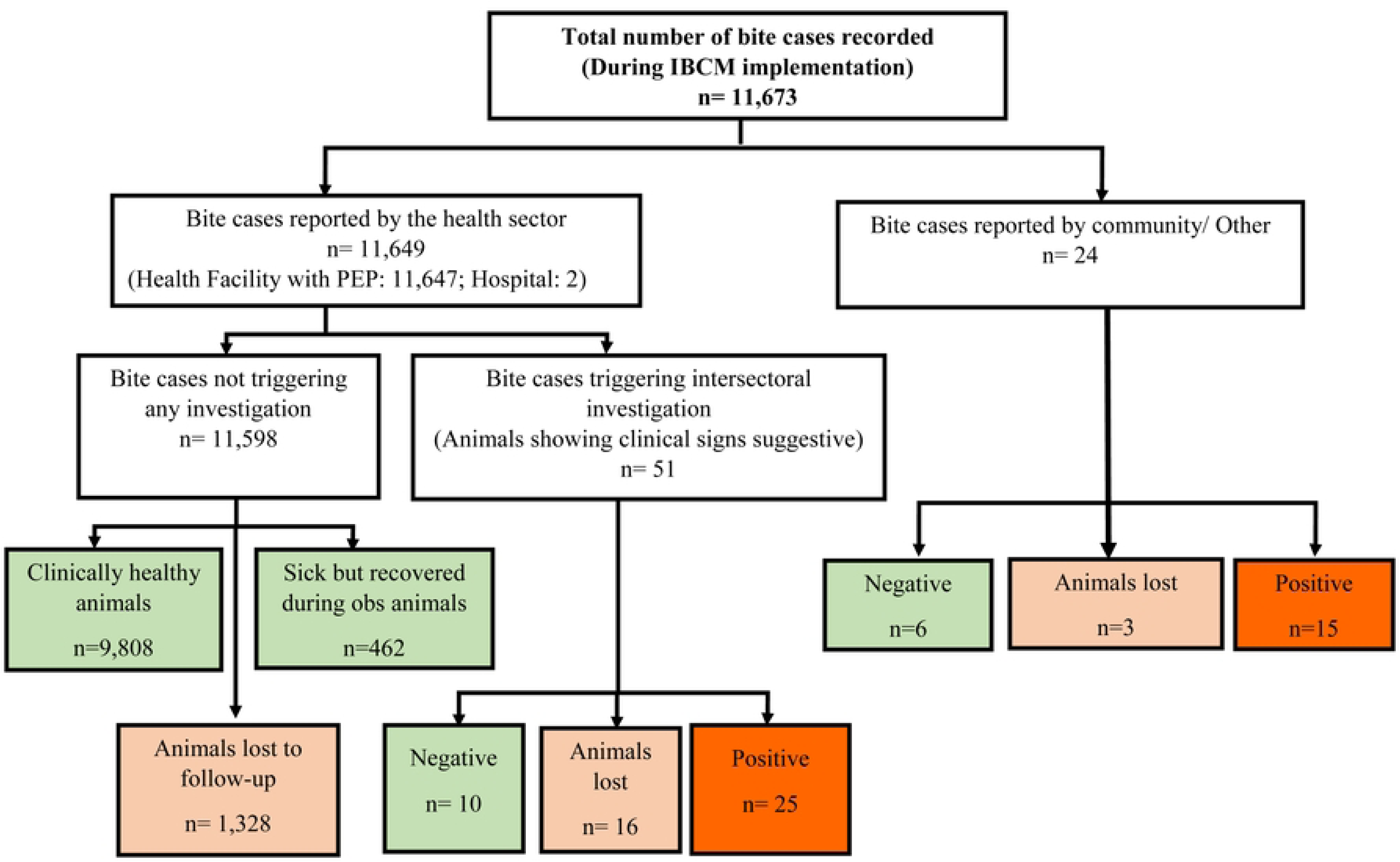
Results of Animal Investigation and Diagnosis under the IBCM Model, September 2024–August 2025 Of the 56 animals sampled, 40 tested positive for rabies virus by RT-PCR, the vast majority of which were dogs (97.5%). None of the animals with positive test results were permanently confined; 25% were free-roaming and 75% were kept under semi-restricted conditions. Most investigated animals were not officially registered with local authorities and had not been vaccinated against rabies within the previous 12 months. At the time of investigation, 24 of the 40 rabies-positive animals had already died, most commonly due to the natural progression of the disease. However, a notable proportion had been killed by community members for consumption. In addition, veterinary services electively euthanised 10 of the 56 animals during the sampling process (**Table 1**).

**Table 1.**
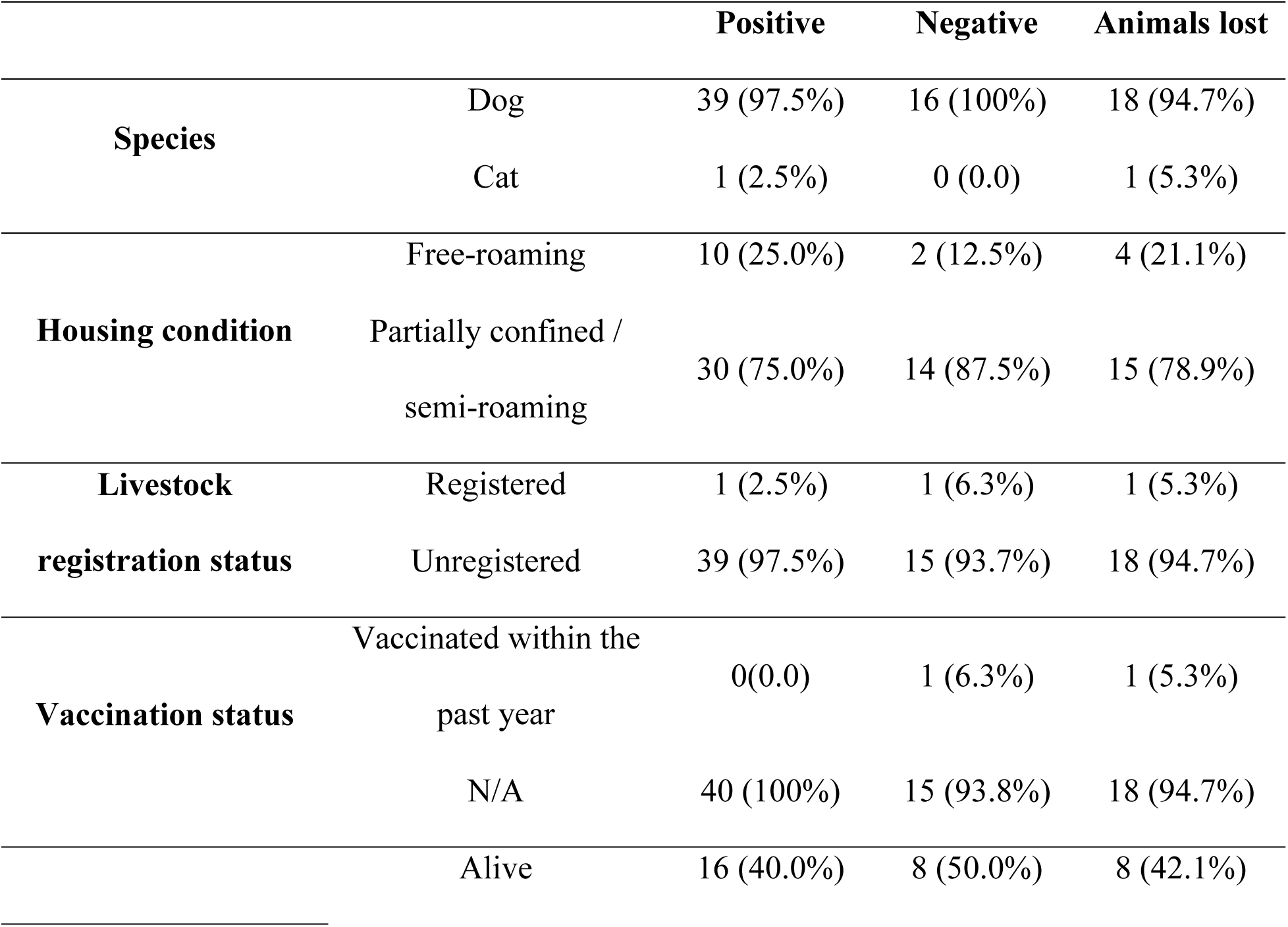
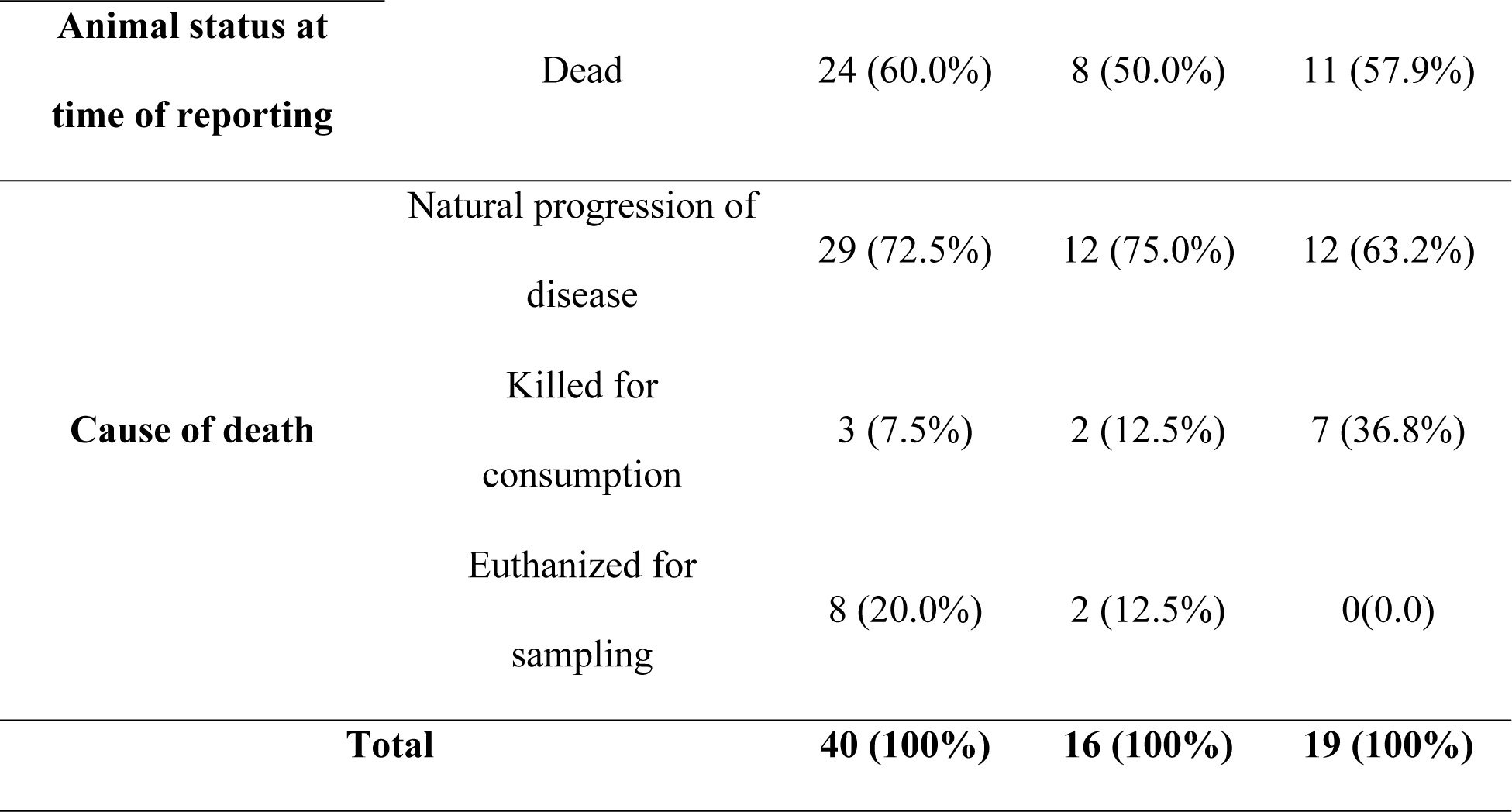
Characteristics of Animals Investigated through the IBCM Model.

A total of 73 animals with observable clinical manifestations were identified through IBCM investigations. Clinical signs could not be assessed in two animals, as they had died some time prior to investigation (these were the animals responsible for bites that subsequently resulted in human rabies deaths). The most frequently documented clinical signs among rabies-suspected animalsanimals included “Biting with no provocation, Altered behavior, and Aggression/irritability.” In contrast, less commonly observed signs included the “dog-sitting” and “licking its own urine” (Fig 4).

**Fig 4.**
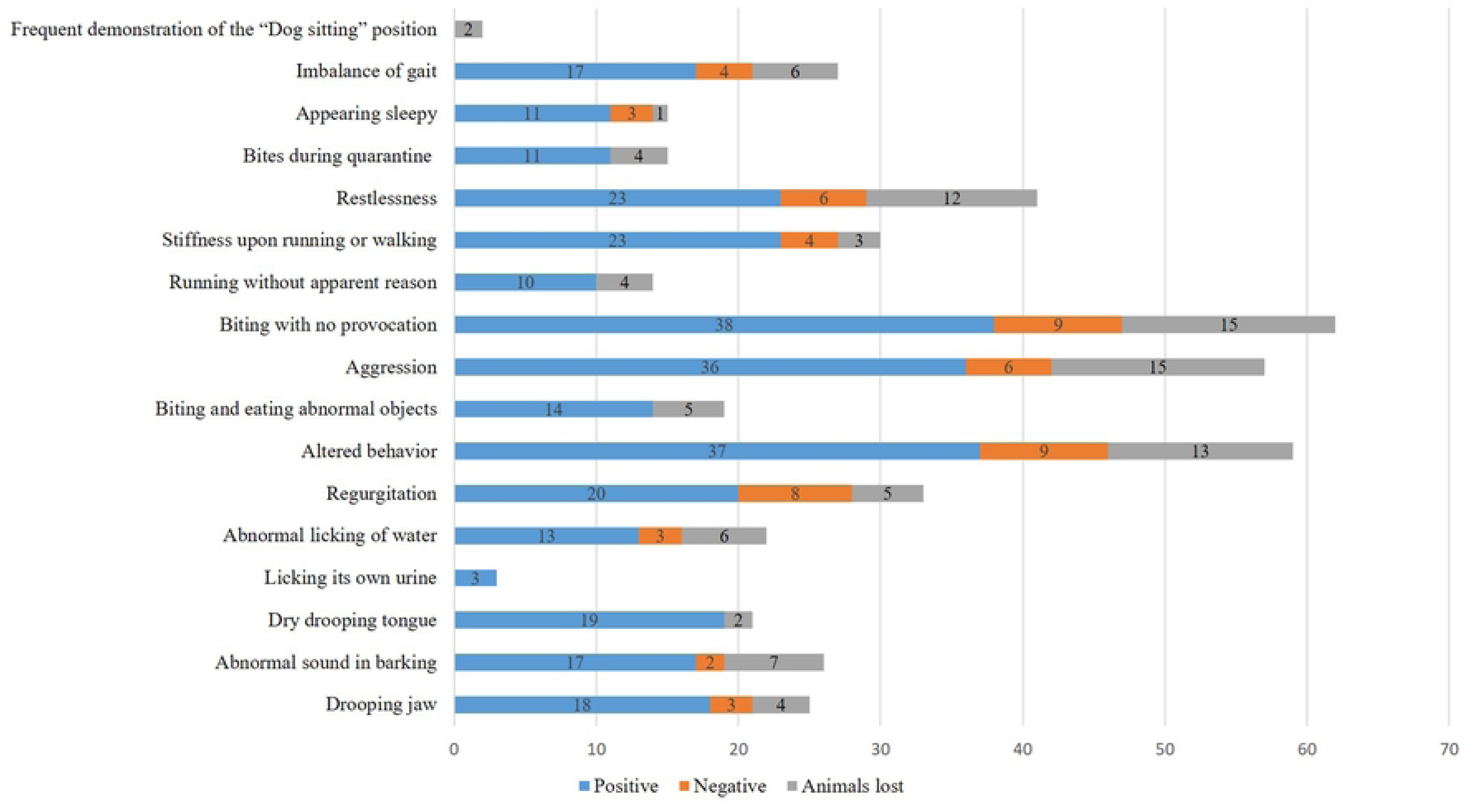
Clinical manifestations observed in rabies-suspect animals investigated under the IBCM model

Figure 5 illustrates the geographic distribution of laboratory-confirmed rabid animals and human bite victims. Rabid animals confirmed by RT-PCR (Fig 5A) were predominantly detected in Dien Ban, Tam Ky, Phu Ninh, Hoi An, Nui Thanh, and Tien Phuoc districts. The mean basic reproduction number (R0) among animals was 1.8, with the number of animals bitten by a single rabid animal ranging from 0 to 7.A total of 83 individuals were identified as having been exposed to animals that tested positive for rabies by RT-PCR, with the highest concentrations recorded in Dien Ban, Dong Giang, Hoi An, Tam Ky, Nui Thanh, Tien Phuoc, Thang Binh, and Que Son districts. On average, each laboratory-confirmed rabid animal bit approximately two people. In addition, 33 individuals were exposed to animals classified as high-risk for rabies (i.e., animals meeting clinical criteria for rabies but for which samples could not be obtained for laboratory confirmation) (Fig 5B).

**Fig 5.**
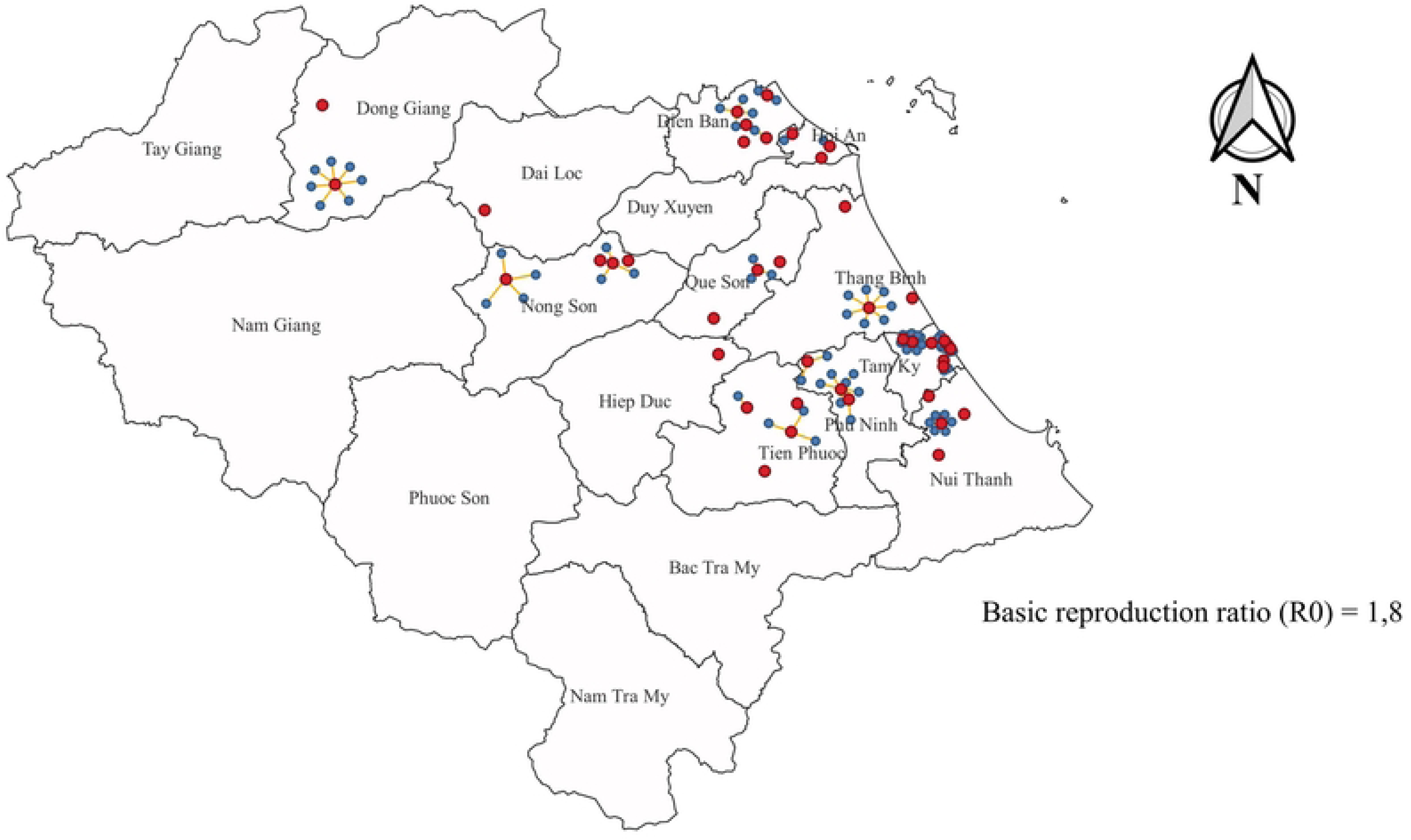
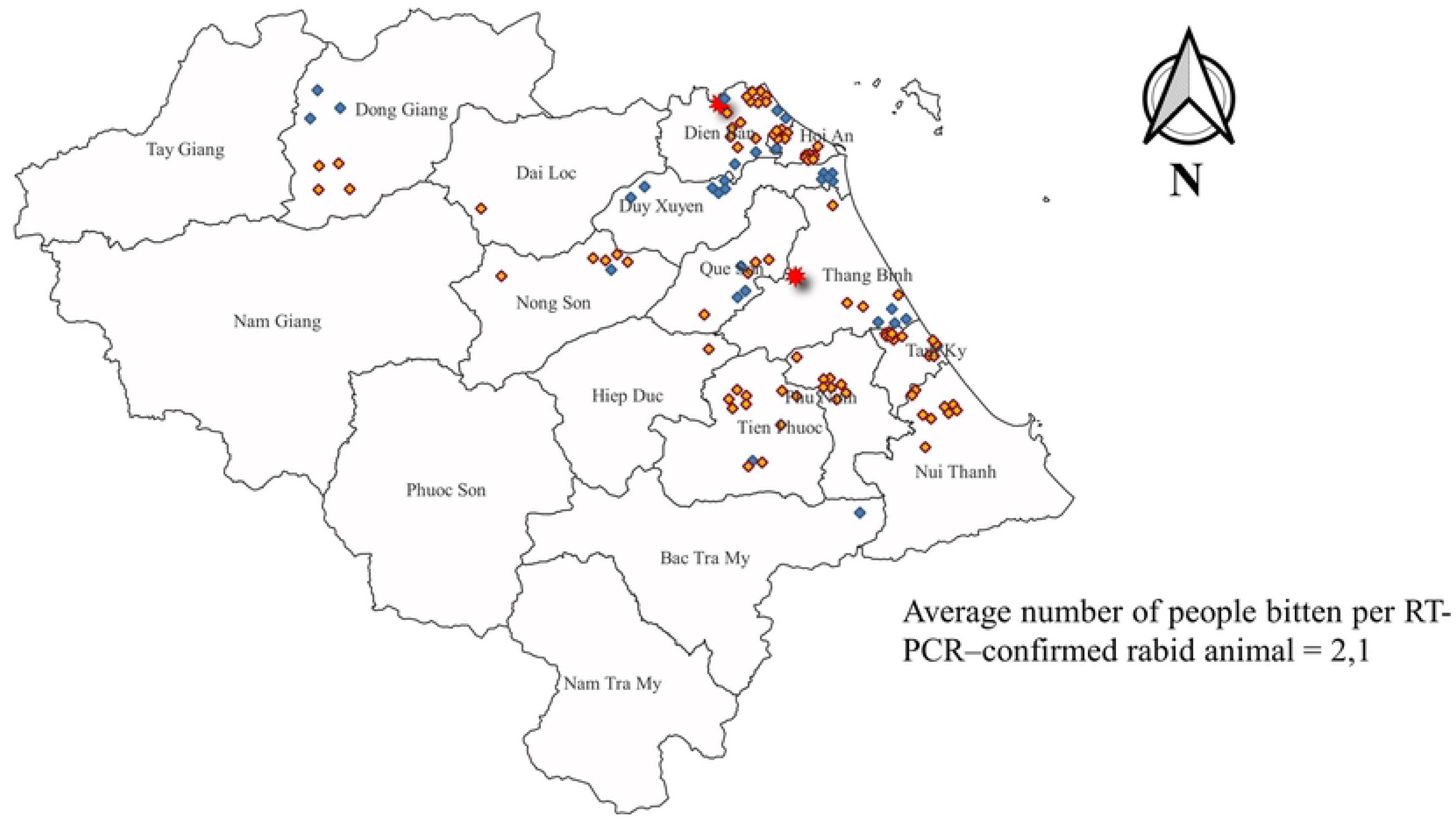
Geographical distribution of rabid animals and human bite victims. Fig 5A. Spatial distribution of laboratory-confirmed rabid animals (red circles) and animals bitten by rabid animals (blue circles); yellow lines indicate transmission links between infected animals. Fig 5B. Spatial distribution of human bite cases, including individuals with confirmed rabies (red stars), individuals bitten by laboratory-confirmed rabid animals (yellow diamonds), and individuals bitten by animals with clinical signs consistent with rabies but without laboratory confirmation due to unavailable specimens (blue diamonds).

The implementation of the IBCM model resulted in significant changes in PEP utilization patterns and exposure risk indicators (**Table 2**). During the intervention period, the average number of individuals receiving PEP at vaccination clinics increased to more than 970 cases per month, compared with an average of 349 cases per month prior to implementation. The proportion of bite victims exposed to high-risk animals who sought PEP also increased from 9.3% to 11.9% following the introduction of IBCM. In addition, the proportion of individuals initiating PEP ≥10 days after exposure rose from 5.9% in the pre-intervention period to 6.6% during the intervention period (p < 0.05).

**Table 2.** Changes in access to PEP and assessment of exposure risk in humans.

|  | Pre-IBCM | Post-IBCM | p-value |
| --- | --- | --- | --- |
| Mean number of PEP treatments per month | 349 | 971 | 0.00* |
| Proportion of bites from high-risk animals | 9.3% | 11.9% | 0.00* |
| Proportion of delayed PEP initiation ($\geq 10$ days after exposure) | 5.9% | 6.6% | 0.00* |

In communes where rabies outbreaks were detected, the average number of registered dogs increased from 507 to 543 dogs per commune within 21 days (**Table 3**). Concurrently, dog vaccination coverage increased from 44.1% to 72.6% during the same period (p < 0.05).

**Table 3.** Changes in dog population estimates and vaccination coverage in areas with detected rabies outbreaks.

|  | At time of laboratory confirmation | 21 days after outbreak detection | p-value |
| --- | --- | --- | --- |
| Mean number of dogs per commune | 507 | 543 | 0.00* |
| Mean dog vaccination coverage per commune | 44.1% | 72.6% | 0.00* |

Knowledge of the primary source of rabies infection among health and veterinary personnel did not show a significant change before and after the intervention (**Table 4**). However, knowledge regarding the diagnosis of rabies in animals among health and veterinary staff increased substantially, from 9.1% prior to the intervention to 55.6% after one year of IBCM implementation, corresponding to an effectiveness increase of 511% (p < 0.05).

**Table 4.** Changes in knowledge of health and veterinary personnel regarding the diagnosis of rabies in animals.

|  | Pre-IBCM |  | Post-IBCM |  | p-value |
| --- | --- | --- | --- | --- | --- |
|  | n | % | n | % |  |
| **Correct knowledge of rabies transmission sources** | 86 | 86.9 | 87 | 87.9 | 0.91* |
| **Good knowledge of animal rabies diagnosis** | 9 | 9.1 | 55 | 55.6 | 0.00* |

\* McNemar test

## Discussion

### Key findings

The IBCM model has been shown to be a suitable surveillance approach in resource-limited settings and where rapid expansion of dog vaccination coverage remains challenging. Previous studies have highlighted IBCM as a representative One Health intervention; however, its implementation, effectiveness, and sustainability may vary depending on local context and available resources [12–14]. The findings of this study demonstrate that the implementation of IBCM using existing local resources is both feasible and effective. During the 12-month implementation period, 75 animals presenting clinical signs consistent with rabies were identified, of which 56 animals were successfully sampled for laboratory testing. Among these, 40 animals (71.4%) tested positive for rabies virus. Despite efforts to collect specimens from all suspected animals, 19 animals could not be sampled, primarily due to carcass disposal by owners (e.g., consumption or discarding into water bodies), which limited the ability to confirm infection status. These findings highlight ongoing challenges in field investigation and the importance of early case detection. Compared with previous surveillance efforts, the IBCM model demonstrated markedly improved effectiveness in identifying rabid animals. Under the routine rabies surveillance programme implemented in Quang Nam Province from 2022 to August 2024 with support from the U.S. Centers for Disease Control and Prevention, 108 animal samples were tested and all were negative for rabies. In contrast, the IBCM model yielded a substantially higher proportion of confirmed cases, indicating superior sensitivity in detecting rabies through targeted, risk-based investigation. Comparable findings have been reported in other settings. In Haiti, approximately 66.0% of animals investigated through IBCM were confirmed rabid, while in Tanzania the proportion was 63.9% [14, 15]. By comparison, in Phu Tho Province, Vietnam, only 32% of animals tested under IBCM were confirmed positive during a 46-month implementation period [10]. The application of structured risk assessment criteria, as recommended by the WHO [11], enabled more efficient identification of high-risk animals and reduced unnecessary testing of low-risk cases. This targeted approach improved the overall efficiency of surveillance activities and optimized the use of limited diagnostic resources, reinforcing the value of IBCM as a practical and scalable One Health strategy for rabies control.

A total of 73 dogs and 2 cats were identified as exhibiting clinical signs consistent with rabies. Among animals confirmed positive by laboratory testing, 97.5% were dogs and 2.5% were cats. This distribution is consistent with findings from previous studies on rabies epidemiology in Africa, Asia, and Vietnam [10, 16, 17]. Animal confinement practices in the study area were generally inadequate. All animals suspected of rabies were either free-roaming or only loosely confined, and the proportion of animals registered with local authorities was very low. None of the laboratory-confirmed rabid animals had documented evidence of prior rabies vaccination. Poor animal management practices have been widely recognized in rabies-endemic settings. In India, for example, a high burden of rabies has been attributed to large populations of free-roaming dogs and insufficient vaccination coverage, making it one of the countries with the highest rabies mortality worldwide [18]. Through the implementation of IBCM, early detection and prompt isolation or removal of suspected rabid animals contributed to interrupting transmission. However, the proportion of animals that were humanely euthanized remained low. This was likely attributable to shortages of trained veterinary personnel at the commune level, a common challenge in Quang Nam, where animal health responsibilities are often undertaken by staff without formal veterinary training. Furthermore, a substantial proportion of animals were lost to follow-up because they were slaughtered for consumption, which poses an additional risk of human exposure to rabies virus [19].

The IBCM model also provided valuable evidence on observable clinical manifestations in rabid animals, as well as on the spatial distribution and transmission dynamics of rabies. In addition to applying exclusion criteria for non-rabid animals, a structured clinical assessment form was used to document observable signs in animals suspected of rabies during field investigations. The most frequently recorded clinical signs were those typically associated with the furious form of rabies, whereas signs characteristic of the paralytic form were observed less frequently. Nevertheless, several less commonly reported but highly suggestive clinical signs—such as drooping of the tongue, inappropriate biting behaviour, and aimless wandering—were also documented. These clinical observations provide important supplementary information for strengthening training programs for veterinary and public health personnel involved in rabies surveillance. The basic reproduction number (R₀) reflects the potential for disease persistence within a population; when R₀ < 1, transmission is expected to decline, whereas R₀ ≥ 1 indicates sustained transmission [20]. In this study, the estimated R₀ was 1.8, with up to seven secondary animal infections attributed to a single rabid dog, indicating a substantial ongoing risk of rabies transmission within the dog population. The magnitude of R₀ may vary according to factors such as dog population density and vaccination coverage. Previous research conducted in Kenya reported an R₀ of 2.44 (95% CI: 1.52–3.36) [21]. The comparatively lower R₀ observed in the present study may reflect the impact of IBCM implementation, particularly through early detection, isolation, and removal of infected animals, thereby limiting onward transmission.

The animal-to-human transmission coefficient observed in this study has limited interpretative value for understanding the overall transmission dynamics, as human-to-human transmission of rabies is exceedingly rare. However, the number of human bite victims remains a meaningful indicator for assessing the likelihood that an implicated animal was rabid. In this study, the animal-to-human transmission ratio was estimated at 2.1, with the highest number of individuals bitten by a single rabid dog reaching seven. This finding is consistent with previous reports, such as that of Xiaoyue Ma et al., which demonstrated that dogs biting two or more people were significantly more likely to be rabid [22]. A substantial increase in the number of individuals seeking PEP was observed following implementation of the IBCM model, with the average monthly number of PEP initiations increasing more than twofold—from 349 cases prior to intervention to 971 cases during the intervention period. This increase likely contributed to a reduction in the number of unprotected exposures to potentially rabid animals, thereby reducing the risk of human rabies deaths. Similar trends have been reported in previous studies. For example, in Haiti, implementation of IBCM resulted in a 30% increase in reported rabies exposure cases in intervention areas between 2013 and 2015 [23], while in Tanzania, Lushasi et al. reported an increase in the average monthly number of bite exposures from 55.7 (range: 15–86) before IBCM implementation to 92.2 (range: 15–174) afterward [14]. The proportion of exposures classified as high risk also increased from 9.3% to 11.9%, which may reflect improved capacity among healthcare workers to correctly identify and classify exposure risk following systematic feedback from animal investigation results. Although the proportion of individuals initiating PEP ≥10 days after exposure increased slightly, the absolute number remained low. This finding should be interpreted with caution, as the observed increase may be influenced by heightened public awareness resulting from extensive dissemination of IBCM-related information through health facilities, veterinary services, local authorities, and mass communication channels, rather than a true delay in healthcare-seeking behavior.

The implementation of IBCM contributed to substantial improvements in outbreak response activities. In communes where rabies outbreaks were detected, the average number of registered dogs increased from 507 to 543 per commune, and vaccination coverage rose from 44.1% to 72.6% within 21 days of outbreak confirmation. These findings indicate that the IBCM framework serves as an effective trigger for targeted vaccination activities and improved dog population registration, both of which are critical components of rabies elimination strategies.

Similar effects have been reported in other settings. For example, a study conducted by Kristyna Rysava in Albay Province, the Philippines, demonstrated that IBCM was an effective tool for identifying areas requiring intensified dog vaccination efforts [13]. Likewise, long-term rabies control programs in Bali, Indonesia, have shown that when high levels of routine dog vaccination cannot be sustained, targeted and rapid vaccination responses guided by IBCM investigations represent an effective alternative strategy. In these settings, vaccination campaigns implemented promptly—often within 10 days following confirmation of a rabid animal—and covering a defined geographic radius around the outbreak have been shown to substantially reduce onward transmission [24].

The IBCM model demonstrated a clear positive impact on improving the diagnostic capacity of health and veterinary personnel in identifying rabies in animals. This finding is consistent with previous studies indicating that IBCM enhances rabies detection through the integration of training, field-based surveillance, and feedback from laboratory-confirmed cases. In Tanzania, Lushasi et al. reported that after 12 months of IBCM implementation, frontline health workers showed substantial improvements in their ability to recognize clinical signs suggestive of rabies. Specifically, all surveyed personnel were able to identify at least three key clinical signs, including hypersalivation (94%), restlessness (86%), unprovoked aggression (86%), and abnormal vocalization (74%), while a smaller proportion also recognized paralysis (34%) and abnormal daytime activity in typically nocturnal wildlife (24%) as indicative of rabies [25]. The observed improvement in knowledge following IBCM implementation can be attributed to several core mechanisms. First, targeted training combined with hands-on field engagement enabled health and veterinary personnel to better recognize both classical and subtle clinical manifestations of rabies in animals. Second, the integration of field investigations with laboratory feedback strengthened experiential learning by linking clinical suspicion with diagnostic confirmation. Third, the increased frequency of exposure to suspected rabies cases—facilitated by systematic reporting from healthcare facilities—provided frontline staff with repeated opportunities to observe, assess, and interpret clinical presentations, thereby enhancing diagnostic confidence and competence over time.

These findings reinforce the role of IBCM as a practical and resource-efficient One Health approach. Rather than establishing parallel or stand-alone surveillance systems, IBCM leverages routine animal bite incidents as a shared entry point for coordinated action between the human and animal health sectors. This study demonstrates that, when integrated into existing local systems, IBCM can substantially enhance the sensitivity of rabies surveillance, accelerate the detection of outbreaks, consequently provide more timely and informed decision-making for post-exposure prophylaxis. Such an approach enables more efficient use of limited resources while strengthening intersectoral collaboration essential for sustainable rabies control.

### Limitations

As a surveillance model primarily reliant on passive reporting, IBCM depends largely on data originating from health facilities providing PEP. Consequently, bite incidents that do not result in healthcare-seeking behavior may not be captured, potentially leading to underestimation of rabies exposure [10]. This limitation was evident in the present study, as two human rabies deaths occurred during the second and third months of implementation, indicating missed opportunities for early detection. Such limitations may be further exacerbated in areas where access to PEP services is limited or absent. Despite these constraints, IBCM has the potential to stimulate broader community awareness and engagement in rabies prevention. This is reflected in the observed increase in PEP utilization in the months following implementation. However, the effectiveness of the model is also highly dependent on human resource capacity. In several communes, veterinary personnel were either insufficient in number or lacked formal training, posing challenges to sustained implementation. Although the IBCM model demonstrated encouraging outcomes within existing resource constraints, strengthening workforce capacity and ensuring adequate staffing levels remain critical to enhancing the long-term effectiveness and sustainability of rabies surveillance and control efforts.

## Conclusion

This study demonstrates that the implementation of the IBCM model within existing health and veterinary systems can substantially strengthen rabies surveillance and response in resource-limited settings. By using animal bite incidents as an entry point for coordinated intersectoral investigation, IBCM enhanced the detection of rabid animals, improved the assessment of human exposure risk, and facilitated more timely outbreak response measures, including increased dog population registration and vaccination coverage in high-risk areas. The model also contributed to meaningful improvements in the knowledge and collaborative practices of health and veterinary personnel, underscoring its value as a capacity-strengthening approach embedded within routine service delivery. Importantly, these gains were achieved without the need to establish parallel systems or introduce substantial additional human resources, demonstrating that effective One Health implementation does not necessarily require new structures but rather better integration of existing ones. Overall, the findings indicate that IBCM represents a practical and scalable strategy for enhancing rabies surveillance and control. By strengthening intersectoral collaboration, improving risk-based decision-making, and optimizing the use of available resources, IBCM can contribute significantly to reducing human rabies deaths. Its broader implementation, supported by standardized operational frameworks and sustained institutional commitment, could play a critical role in advancing the goal of eliminating dog-mediated human rabies by 2030.

## Data Availability

All relevant data underlying the findings of this study are included within the manuscript and its Supporting Information files

## Acknowledgements

The authors would like to express their sincere gratitude to the institutions and individuals who played an essential role in the implementation and support of the IBCM programme in Quang Nam Province. We gratefully acknowledge the National Institute of Hygiene and Epidemiology, the Pasteur Institute in Nha Trang, the Da Nang Center for Disease Control, the Da Nang Department of Health, and the Da Nang Department of Agriculture and Environment for their collaboration and support. We also extend our sincere thanks to Dr. Kiem Van Tran, Dr. Vinh Dai Nguyen, and Dr. Quang Cong Huynh for their valuable contributions to this work.

